# Reasons for Protocol Deviations & Missed Antibiotic Doses in Patients Undergoing Oncological Endoprosthetic Reconstruction

**DOI:** 10.1101/2024.05.06.24306755

**Authors:** Serena Uppal, Hadia Farrukh, Michelle Ghert, the PARITY Investigators

## Abstract

**Background:** Research study protocols are critical in study design, however study implementation can be subject to protocol deviations. The Prophylactic Antibiotic Regimens in Tumor Surgery (PARITY) trial assessed prophylactic intravenous antibiotics on the rate of surgical site infection in patients undergoing oncological endoprosthetic reconstruction of the lower extremity. The objective of this study was to identify the protocol deviations – missed antibiotic doses – in the PARITY trial, and determine the causes of the protocol deviations.

**Methods:** This study is a secondary analysis of the PARITY trial data set. The patients in the original trial were randomized to receive either 24 hours or five days of postoperative intravenous antibiotics, for a total of 15 doses. Patients that missed doses and the reason for each missed dose were recorded in the database and summarized descriptively herein. This data was then compared between clinical sites with high and low volume, between nationalities and between the economic development status of the clinical site location.

**Results:** The PARITY trial included 604 participants with 218 patients missing at least one antibiotic dose. The most common reason across all clinical sites was that patients were discharged earlier than the five-day protocol. This finding was consistent across high and low volume sites and across clinical sites in areas of different economic development. The reasons for protocol deviations varied across clinical sites of different nationalities.

**Conclusions:** Protocol deviations – missed antibiotic doses – were common but of minimal severity in the PARITY trial. The most commonly reported reason for missed antibiotic doses was discharge earlier than the five-day protocol and not due to clinical site personnel error. Nevertheless, this study did identify actionable improvements to study protocol adherence such as careful monitoring of drug administration, investigator equipment, and availability of study drug.

---

Protocols are essential for experimental research and are a cornerstone for clinical practice. Oftentimes, especially in large scale studies, errors can occur in following the protocol. This is referred to as a protocol deviation. It is important to note that a deviation is not a violation which is infinitely more significant (Bhatt, 2012). Deviations are common and do not challenge the validity or existence of the study, while violations can completely derail or discontinue the study. This is not to say that studies are completely unaffected by protocol deviations, they still need to be reported in charts and avoided when possible. An example of a protocol deviation, that will be heavily considered in this analysis, includes the failure of clinical sites to administer the correct number of antibiotic doses.

Protocol deviations were a significant factor in the original PARITY trial. Missed antibiotic doses among patients throughout all the sites could have impacted the data in unforeseen ways. In a previous analysis, factors such as site location, site volume, and development status were considered and evaluated to determine possible trends surrounding missed doses. Site location considered protocol deviations among each country and continent, while site volume categorized each site based on the median of six patients, as either low volume (less than or equal to six patients) or high volume (greater than six patients). The development status for each site was split between developing and developed countries.

The protocol stated that all patients should receive three antibiotic doses on the first postoperative day. The patients in the treatment group should continue to receive three antibiotic doses per day over the next four days. The patients in the placebo groups should also continue to receive three doses per day over the next four days, however they will be injected with the placebo instead of the antibiotic. The protocol deviations, in this case missed antibiotic doses, were split into either unblinded or blinded doses. Each patient was set to receive three antibiotic doses on the first day regardless of the assigned treatment regimen, therefore the doses delivered on the first day were unblinded. For the following four days, all 12 doses were blinded for administrators and patients as both were made unaware if it was the antibiotic or the placebo.

The results from this previous evaluation indicated a variety of trends amongst all the categories examined. Low volume sites missed 7.2% of unblinded doses, while high volume sites only missed 3% of unblinded doses. There was minimal difference between low and high volume sites for the percentage of missed blinded doses. Sites in Austria missed the most unblinded doses (33.3%), while sites in the United States missed the most blinded doses (15.4%). European sites missed the most unblinded doses (7.8%), while North American sites missed the most blinded doses (13.5%). Developed countries missed more doses for both unblinded (4.6%) and blinded (12.6%) compared to developing countries who only missed 0.7% of unblinded doses and 8.6% of blinded doses. A statistical test needs to be conducted for each category to determine the significance of these results.

## Materials and Methods

### The PARITY Trial

The Prophylactic Antibiotic Regimens in Tumor Surgery (PARITY) trial was a multicenter international blinded randomized controlled trial. The data was collected from 48 clinical sites in 12 countries with a total of 604 participating patients (PARITY Investigators et al., 2022). The patients who were treated and qualified for this study had undergone limb reconstruction surgery as a treatment for osteosarcoma, oligometastatic bone disease, or soft tissue sarcoma. The project compared a one-day versus five-day regimen of postoperative intravenous antibiotics to determine which option was more effective in reducing the extent of surgical site infection (PARITY Investigators et al., 2022). This study is a secondary analysis of the previously conducted PARITY trial. Protocol deviations throughout the trial, in the form of missed antibiotic doses, were analyzed.

### Data Source

All relevant data was obtained from the original PARITY database. The specific variables that were considered were site number, site location, patient identification number, number of administered antibiotic doses for each patient, and staff notes which indicated the reason for the missed dose.

### Data Analysis

Due to the nature of the data, a descriptive analysis was performed. Microsoft Excel was used to sort and organize the variables in order to observe trends. Each patient has an identification number that indicates site number and location. In the database, the total number of antibiotic doses administered to each patient was recorded and corresponds to each identification number. If patients were subject to protocol deviations (missed antibiotic doses), the reasons for this were indicated in the database. Each patient who missed at least one dose was sorted into one of the following groups: no reason indicated, drug unavailable, staff error, discharged early, patient refusal, patient transferred, additional/prolonged surgery, intravenous (IV) issues, complications/new onset of disease, death, allergy, and staff determined two doses were sufficient. This data was then combined with the previous analysis that considered site location, site volume, and development status. The most common reason for missed doses was determined for each country, each continent, low volume sites, high volume sites, developing locations, and developed locations.

## Results

### Study Population & Relevant Data

The original PARITY trial included 604 patients and the data for each patient was considered in this secondary analysis. However, only 218 patients missed at least one antibiotic dose and were subject to protocol deviations, therefore these participants were the only relevant data points for this study.

### Reasons for Protocol Deviations

The administrators of each site were required to report the reason for the protocol deviation for every patient who missed antibiotic doses. This data was compiled for all 218 patients and sorted into various groups.

Using the number of patients that missed doses for each reason (N) and the total number of patients that missed doses (218), the percentage for each reason was calculated. The findings are summarized in Table I.

**Table I:** Reasons for Protocol Deviations.

| **Reason For Missed Antibiotic Doses** | **Number of Patients that Missed Doses for this Reason (N)** | **Percentage of Total Number of Patients that Missed Doses ((N/218)\*100)** |
| --- | --- | --- |
| **Additional/Prolonged Surgery** | 5 | 2.3% |
| **Allergy** | 1 | 0.5% |
| **Complications (e.g. onset of new disease)** | 10 | 4.6% |
| **Death** | 2 | 0.9% |
| **Discharged Early** | 126 | 57.8% |
| **Drug Unavailable** | 2 | 0.9% |
| **Intravenous (IV) Line Complications** | 3 | 1.4% |
| **No Reason Indicated** | 29 | 13.3% |
| **Patient Refused** | 9 | 4.1% |
| **Patient Transferred** | 2 | 0.9% |
| **Staff Error** | 23 | 10.6% |
| **2 Doses Sufficient (Skipped 3rd on Purpose)** | 6 | 2.8% |

### Cross Reference with Factors Contributing to Protocol Deviations

Using the data from the previous analysis of factors contributing to protocol deviations, Table I, and the PARITY data set, site volume, site location, and development status were cross referenced with the reasons for missed antibiotic doses. A summary of these findings are represented in Tables II, III, and IV.

**Table II:** Site Volume Cross Reference with Reasons for Missed Doses.

| **Site Volume** | **Most Common Reason for Missed Doses** | **Total Number of Patients that Missed Doses For Most Common Reason (N)** | **Total Number of Patients that Missed Doses (M)** | **Percentage of Most Common Reason ((N/M)\*100)** |
| --- | --- | --- | --- | --- |
| **Low Volume (Less Than/Equal to 6) Sites** | Discharged Early | 14 | 35 | 40% |
| **High Volume (Greater Than 6) Sites** | Discharged Early | 112 | 183 | 61.2% |

**Table III:**
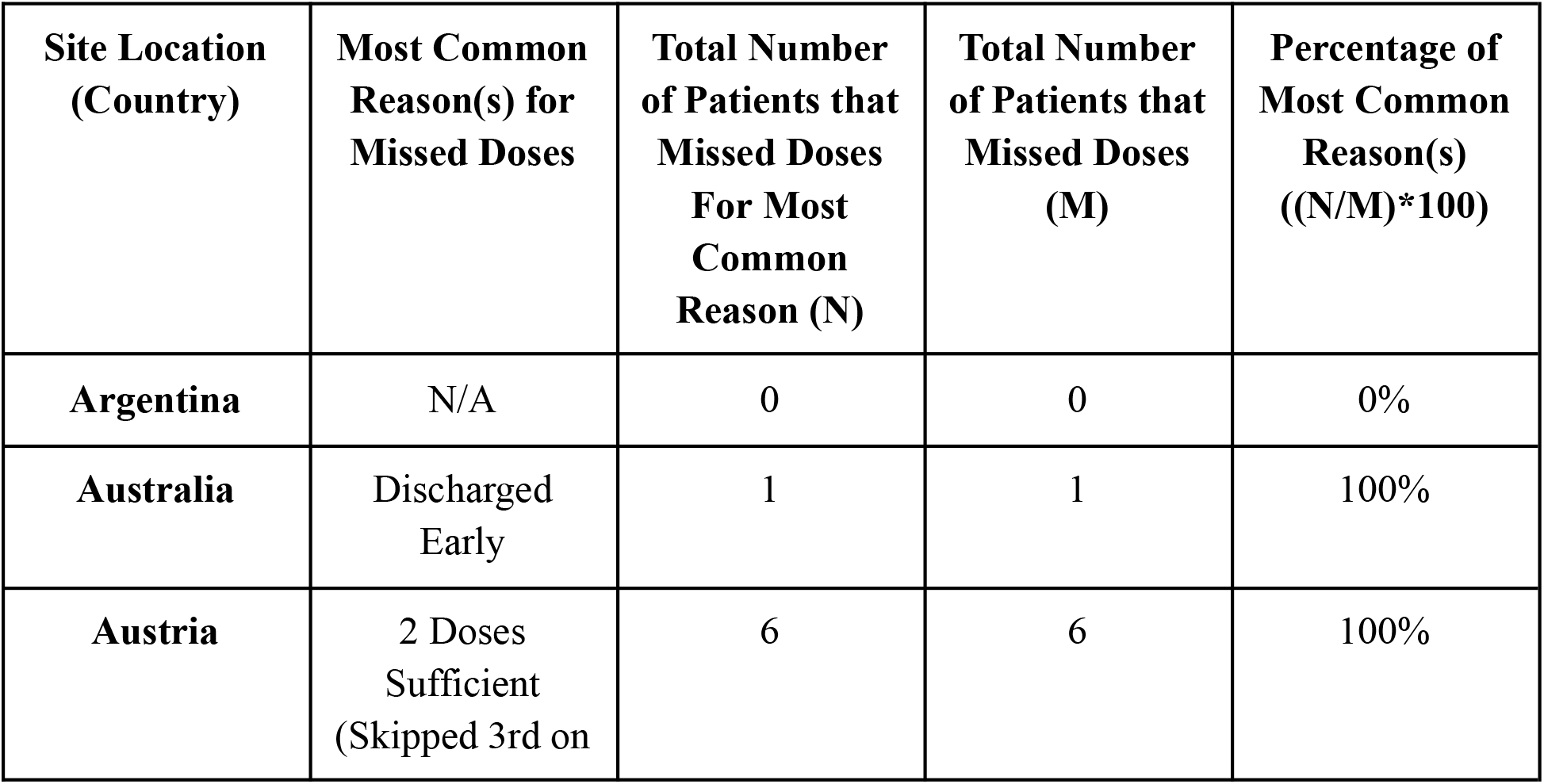
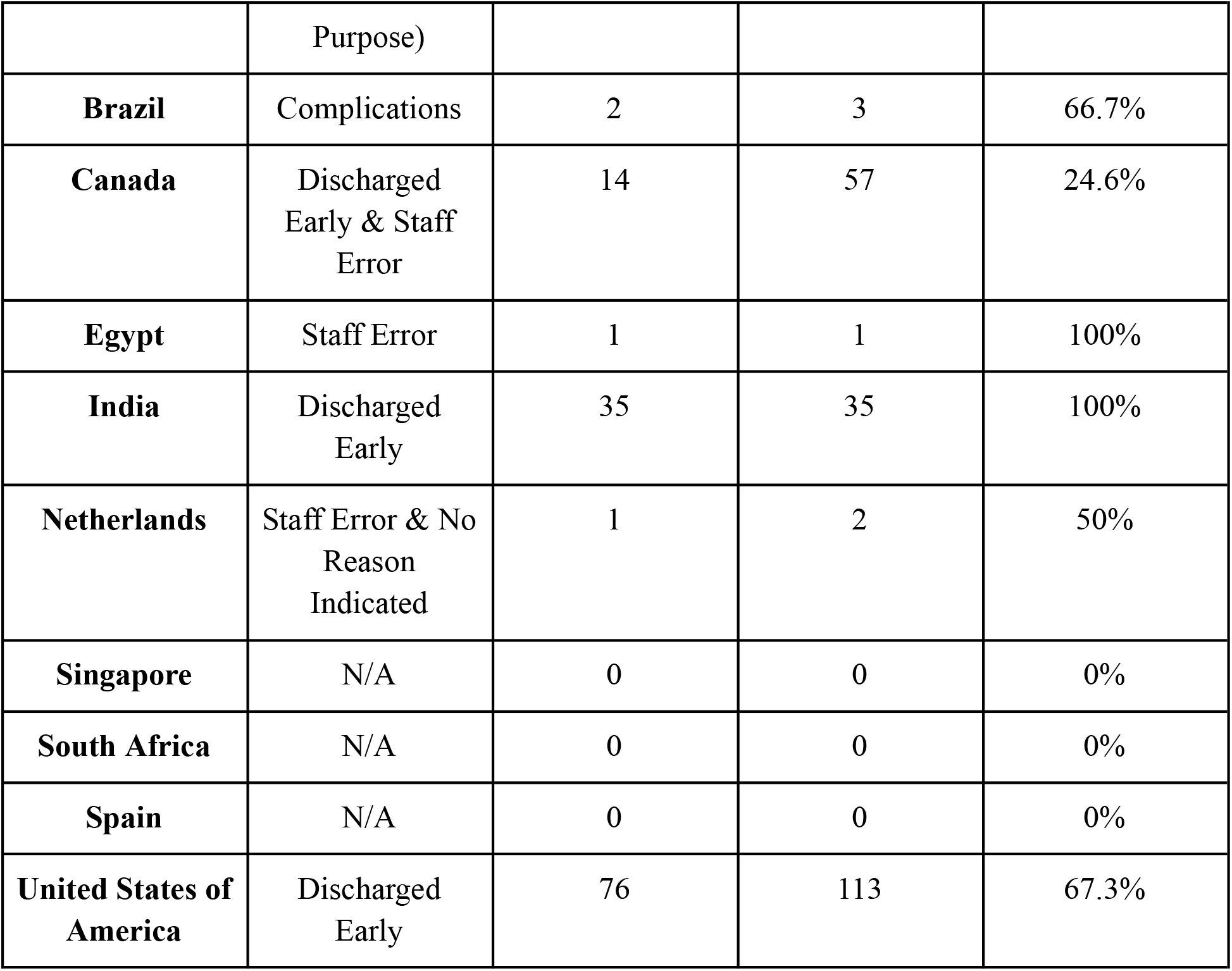
Site Location Cross Reference with Reasons for Missed Doses.

**Table IV:** Development Status Cross Reference with Reasons for Missed Doses.

| **Development**<br>**Status** | **Most Common**<br>**Reason for**<br>**Missed Doses** | **Total Number**<br>**of Patients that**<br>**Missed Doses**<br>**For Most**<br>**Common**<br>**Reason (N)** | **Total Number**<br>**of Patients that**<br>**Missed Doses**<br>**(M)** | **Percentage of**<br>**Most Common**<br>**Reason**<br>**((N/M)\*100)** |
| --- | --- | --- | --- | --- |
| **Developing** | Discharged<br>Early | 35 | 39 | 89.7% |
| **Developed** | Discharged<br>Early | 91 | 179 | 50.8% |

In Table II, the analysis considered low and high volume sites. The most common reason for missed doses was determined by calculating which reason encompassed the most patients in each category. The percentage was calculated using the total number of patients that missed doses for the most common reason (N) and the overall total number of patients that missed doses (M).

Table II indicates that patients being discharged early was the most common reason among both low and high volume sites. The majority of patients that missed doses in high volume sites (61.2%) were due to being discharged early.

Table III considers each participating country in the original PARITY trial. The percentage indicating the prevalence of the most common reason within each country was calculated using the total number of patients that missed doses for that reason (N) and the total number of patients that missed doses for any reason in the corresponding country (M). Table III indicates that Argentina, Singapore, South Africa, and Spain did not miss any doses. Patients in Australia and India that missed doses were all due to being discharged early. All patients in Austria missed doses due to the belief that two doses were sufficient and they did not need to be injected with the third, mandatory dose. The only patient that missed doses in Egypt was a result of staff error. The majority of missed doses in Brazil were due to complications, and the majority of missed doses in the United States of America was due to patients being discharged early. Canada and the Netherlands both had multiple reasons for missed doses since the same number of patients missed doses for each reason. Canada had 14 patients that missed doses due to being discharged early and staff error. A similar case occurred in the Netherlands, however the most common reasons for missed doses were staff error and no reason indicated.

Table IV calculates the percentage of each common reason using the total number of patients that missed doses for the reason indicated (N) and the total number of patients that missed doses for each development status (M). The data shows that the most common reason among both developing and developed sites was patients being discharged early. The majority of missed doses for both development statuses were due to this reason.

The purpose of cross referencing site data, based on specific characteristics, with the reasons indicated for missed antibiotic doses, based on patient chart data, was to determine the most common reasons for protocol deviations on a smaller scale. Table I represents the reasons for protocol deviations among the entire 218 patients that missed doses, while Tables II, III, and IV consider various factors and the traits that define each site.

## Discussion

To our current knowledge, this is one of the first studies to consider the reasoning and demographics of protocol deviations in an international, randomized control trial. The results suggest that patients undergoing oncological endoprosthetic reconstruction are subject to altered treatment methods due to the deviation and errors from their medical team. Descriptive statistics used in the analysis support this theory and indicate that regardless of the effort and detail that is implemented in developing protocols for trials, deviations are inevitable. Seemingly, this is unsurprisingly due to the scale of the original PARITY trial which involved the coordination of 48 different sites, with varying volumes, around the world.

For the purpose of full transparency, all of the indicated reasons from the patients charts were included in the results. However, it is important to consider that some of the reasons indicated are outside the control of the site and therefore not the fault of the administrators. Examples of this, that cannot be considered a valid error or deviation, include being discharged early, when the patient refuses treatment, when the patient is transferred, if the patient requires additional or prolonged surgery, if complications arise, if the patient dies, and if the patient is allergic to the antibiotic treatment. Factors that can be considered valid protocol deviations or errors and require improvement in future trials include when there was no reason indicated, the drug was unavailable, staff error, IV issues, and when the site determined that, according to their standards, only two doses were needed to sufficiently recover. The analysis of this data is heavily impacted by this revelation and the methods in which the results were interpreted.

As suggested by Table I, the most common reasons for patients to miss doses and deviate from the protocol include being discharged early (57.8%), staff errors (10.6%), and for no apparent reason (13.3%). Based on the previous considerations, patients discharged early are not a deviation from the protocol. Table II and IV are consistent with the results from Table I. The most common reason for all factors regardless of site volume or development status was patients being discharged early. Table III decreases the scope of the analysis by narrowing the data and site to its residing country. This provided increased variation in the results with different countries providing reasons that do not include patients being discharged early. Austria, Egypt, and the Netherlands are all examples of countries that were subject to valid protocol deviations based on our previous definition. The findings from this analysis suggest that as an overall experiment, considering all patients regardless of grouping, varying site volumes, and varying development statuses, the PARITY trial sufficiently administered doses with minimal protocol deviations, as the majority of protocol deviations in each case occurred due to the patient being discharged early.

### Previous Literature

Findings from previous studies analyzing protocol deviations among various trials have yielded similar results to this analysis. A recent study conducted by Ghooi et al. (2016) assessed the protocol deviations present in clinical trials. Using a grading system from 1-5, the effects of protocol deviations were categorized with Grade 1 having no effect on the patients or data to Grade 5 being the most extreme classification with the subject or patient dying as a result of the deviation (Ghooi et al., 2016). The analysis concluded that the majority of deviations fell under Grades 1 or 2 with 289 of the total 368 deviations being classified in both categories combined. Zero deviations were recorded as Grade 5 over the three year period. Additionally, it was determined that when the deviations were split into either minor or major, there were substantially less major deviations and the total number was decreasing throughout the years (Ghooi et al., 2016). The results from this analysis are consistent and in line with the data from the secondary analysis of the PARITY trial. Although, in both cases, the protocol deviations were minor and did not appear to affect either of the trials, it is still paramount that they are reported and considered in the overall analysis. The study conducted in 2016 and this current analysis are comparable and follow similar methods including the use of Excel for data analysis.

As mentioned previously, there is limited research on the specific reasons behind protocol deviations. This analysis is unique in determining the most common reasons for missed antibiotic doses. The study conducted by Ghooi et al. (2016) broadly discussed examples of deviations, but they did not conduct further analysis on the reasoning. Instead, they classified the deviations based on severity. However, this can still be compared to the current findings from this secondary analysis, since the reasons for protocol deviations can also be categorized. One of the causes of protocol deviations that Ghooi et al. (2016) discussed involved the globalization of clinical research. Since data is emerging from various countries, it is impossible to regulate or ensure the quality is consistent on a global scale. This can lead to some discrepancies in the results that may affect the conclusions drawn by the study. Comparing these findings to Ghooi et al. (2016) provides a basis in understanding the data, which allows for further analysis to be conducted. The reasoning for protocol deviations, noted in patient chart data, can help to provide clarity and aid in determining the exact problem that should be avoided in future protocols.

Previous analyses have conducted statistical testing and various procedures to develop conclusions on protocol deviations, in order to aid in future protocol development. Nettleman & Bock (1996) determined that there was an increased chance of missing doses with an increased amount of required doses per patient. This is applicable and can be seen in the original PARITY trial, where a considerable amount of doses needed to be administered throughout the experiment. In the future, to avoid missing antibiotic doses, the number of daily required doses can be altered. It is evident that protocols in future trials must determine how many doses are necessary. If the results will not be affected, trials may benefit from less required doses. The findings from Coleman et al. (2013) build on the current study and consider measures that can be put in place to help reduce protocol deviations. Doses were significantly less likely to be missed with the use of clinical dashboards, Root Cause Analysis (RCA) meetings, and the publication of a National Patient Safety Agency (NPSA) Rapid Response Alert (Coleman et al., 2013). Clinical dashboards report antibiotic dose administration and indicate overdue doses, while RCA and NPSA meetings discuss action plans to reduce the omission of antibiotics ensuring that every patient receives their required doses. These techniques can be attributed to the original PARITY trial and future studies to further minimize protocol deviations.

### Implications

This secondary analysis of the PARITY trial suggests that although valid protocol deviations were minimal, there were still a non-zero amount of errors conducted on the majority of clinical sites. In order to maintain the integrity of the research and results from the original trial, the protocol must be followed correctly. Additionally, this can be applied to any and all protocols in experimental research. The protocol cannot be deviated from, otherwise the results are in jeopardy or could render invalid. Using the results, future protocols can extrapolate this data and apply it to their own, in order to avoid similar errors.

### Strengths & Limitations

The present study had several strengths. The most notable one being the multi-centered approach by the original PARITY trial. This allowed for a large, diverse sample size. The database provided a multitude of information on each site including volume, location, and the number of administered doses. This secondary analysis took advantage of the large data set to provide the largest scope possible for each factor. The analysis options were exhausted to determine every possible relationship present in the data set.

Although there were many strengths of this study, there were also some limitations. Due to the execution of the PARITY trial on a substantially large scale, it was impossible to ensure consistency in discharging patients. The protocol stated that each patient should receive antibiotic treatment over a five-day period, but each location followed their own site-specific procedures. It was impossible for this study to standardize the length of patient stays due to the varying out-patient procedures that are in place around the world. Therefore, this ultimately impacted the secondary analysis with the majority of patients being released and discharged earlier than expected.

## Conclusion

The reasons for protocol deviations were consistent among all sites, despite their volume or development status. The most common reason that patients missed doses was due to being discharged from the site earlier than anticipated. The most common reasons that are considered errors, as the trial had no control over patient length of stay at each site, were due to staff error and no reason indicated. Protocol deviations, in this case and in other studies, have been minimal and do not appear to impact the overall study, however it is extremely important to continue to report deviations in order to maintain the integrity of the experiment, ensure reproducibility, and to uphold the health and safety of the participants. Protocol deviations can appear differently across various trials, but they can be grouped together when considering a solution. Minimizing the reasons indicated in this analysis, now that the causes are known, and implementing the strategies discussed can improve protocols and the methods in which studies are conducted to avoid deviations in the future.

## Data Availability

All data produced in the present study are available upon reasonable request to the authors

## Appendix

### Appendix A. Reasons for Protocol Deviation

Additional/Prolonged Surgery: Patient was subject to additional surgery and had to return to the operating room.

Allergy: Patient allergic to antibiotic dose.

Complications: New onset of disease, concerns about patient safety, complications in surgery, deteriorating condition etc.

Death: Patient passed away.

Discharged Early: Patient discharged from hospital before the full five-day period.

Drug Unavailable: Drug unavailable at the site.

Intravenous (IV) Line Complications: Inability to inject antibiotics due to complications with IV.

No Reason Indicated: The site did not indicate the reason that the dose was missed.

Patient Refused: Patient retracted their consent and refused the dose.

Patient Transferred: Patient transferred, miscommunications due to transfer.

Staff Error: Nurse error or forgot to administer dose.

2 Doses Sufficient: Site determined that 2 doses were all the patient needed and skipped the third dose on purpose.

